# A Simple Method of Finding an Approximate Pattern of the COVID-19 Spread

**DOI:** 10.1101/2020.05.24.20112292

**Authors:** Hemanta Kumar Baruah

**Affiliations:** Department of Mathematics, The Assam Royal Global University, Guwahati, Assam - 781035, INDIA

**Keywords:** Corona virus disease, country, pandemic, short term forecasting

## Abstract

We are going to show that the pattern of spread of COVID-19 outside China is not monotonic. We have considered the data outside China because we are going to study the data starting from March 21, and by that time the spread had almost come to a stop in China. We have used for our analysis data on total cases outside China till April 25, 2020, and data from April 26 to April 30 for comparison of forecasts and observed values. Right from the beginning the spread pattern was nonlinear, and by the end of the third week of March the nonlinearity became nearly exponential. The exponential pattern thereafter has changed by around March 28, April 5, April 11 and April 18. Since March 21, the spread is following a nearly exponential pattern of growth changing observably at almost regular intervals of seven days. It is but natural that at some point of time the countries that had been contributing in observably large numbers to the total cases would start to show diminishing growth patterns. Therefore long term forecasts using our method would give us slightly overestimated results. However, for short term forecasting our simple method does work very well when we consider the total number of cases in the world and not in any particular country.

## INTRODUCTION

The objective of our work is to find out in a *very simple way* an approximate pattern explainable mathematically of the spread of the corona virus disease that has affected the entire world in just five months. We have taken the data for our analysis from Worldometers.info [1] published on April 25, 2020. It is to be noted that the data regarding the COVID-19 matters made available by Worldometers.info are regularly edited to make small changes in the data published a few days ago. As such, the Worldometers.info data published on April 25 would be very slightly different from that published on April 18, say. To get a proper picture of the spread pattern, we have considered the case of COVID-19 spread outside China because as per the Worldometers.info data, in China the spread has almost come to a halt already. Such data may be underestimations of the real figures because there would anyway be unreported cases. However, to get an idea regarding the spread pattern, small aberrations are not quite relevant because by this time the total number of cases registered till any particular day is observably very large.

Researchers have used data from Worldometers.info regarding COVOD-19 matters for mathematical modeling of the spread pattern (see for example [2]). In that article published on 15 April, 2020, the data analyzed were those that were available till the end of March, 2020. Not much of data was available at that time so as to get a clear picture of the spread pattern.

Kucharski *et. al*. [3] attempted to estimate the dynamics of transmission of the disease when it was in the very early stage in February, 2020. They used a geometric random walk process and went for Monte Carlo simulation for inferences. Regarding estimation of the spread pattern of the pandemic, Gondauri *et. al*. [4] concluded that it is difficult to predict about the spread as on March 30. The data that they have used were of the initial period of the outbreak. A clearly observable pattern had not emerged by that time. Wu *et. al* [5] studied the spread pattern of the outbreak during that initial period. They used a susceptible-exposed-infectious-recovered model to simulate the epidemic right when it was in the very initial stage in Wuhan City of China. Though not spread rates, Baud *et. al*. [6] and Wang *et. al*. [7] studied mortality rates caused by the virus.

We have observed that the data of the initial period of the outbreak were not satisfactory enough for extrapolation because no specific pattern was reflected in the data at the initial stage. From the data on total number of COVID-19 positive cases till any particular day, we have further observed that constructing a mathematical model using a *monotone* increasing function of time is of no use, because the spread pattern is not monotonic. From January 22, 2020, onwards COVID-19 positive cases started to appear outside China. Within a week or so, it started to be nonlinear. By the end of the first week of March, it started to become very highly nonlinear. By around the third week of March, it started to become nearly exponential.

In what follows, we are going to study the character of the spread pattern using classical mathematical procedures. We are not really going to fit a curve in the sense of mathematical modeling. Our objective is to show that currently the spread pattern is not a monotone function of time. We have found that the spread is nearly exponential with the pattern changing at almost regular intervals of seven days.

Finally, we have attempted to forecast the total number of cases from April 27 to April 30. We have found that extrapolation using our procedure does return values that are quite close to the observed data during this period.

## METHODOLOGY

We shall begin with the following simple explanation. Let *y* be the value of total number of COVID-19 cases on any given time *t*. If *y* is a function of *t* following

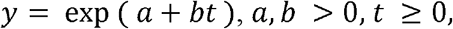

where *a* and *b* are constants, then

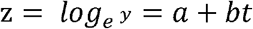

is linear in *t*. For such a *y* following the exponential pattern, the rate of change of *y* is also exponential. Now as (*a* + *bt*) is a first degree expression, the first order differences Δ*z* of *z* would be constant for equidistant *t*. Therefore if we observe that the first order differences of observed values of z for equidistant values of *t* are very nearly constant, we can conclude that *y* is very nearly exponentially increasing.

The COVID-19 data published by Worldometers.info in the form of graphs are based on cumulative daily data depicted in such a way that the numerical values can be found from the graph directly. We have therefore made an attempt to see approximately from which date onwards the values of z started to become approximately linear in time. To do that we would need to look at the time series data downwards and this is where our approach is different and simpler than time series analysis.

From the data it is evident that at the initial stage, the spread function was not exactly of the exponential type. However, even from day-1, January 22, 2020, the total number *y* of cases was nonlinear already, because Δ*y* was far from constant. In other words, it was nonlinear in nature right from the beginning. But it was not of the exponential type till the end of the third week of March. Nonlinearity increased thereafter and ultimately it is nearly exponential now.

## ANALYSIS

We shall study the total number of COVID-19 positive cases outside China starting from April 25 downwards. Total cases here refer to total cumulative count, and this figure includes deaths, active cases and discharged patients. We would like to reiterate that the data regarding the COVID-19 matters made available by Worldometers.info are regularly edited to make small changes in the data published a few days earlier, and therefore the readers may find some small amounts of changes in the total number of cases *y* shown in the tables below. However, the changes are too small in comparison to the largeness of the values of *y*.

In the five tables below we shall depict the total number of cases from April 25 to March 21 downwards. We shall explain with the help of the tables why we have mentioned that the spread function is not monotonic. From the tables, we can see that the values of z are very nearly linear, linearity being reflected by approximate constancy of the values of Δ*z*.

From Table-1, we can observe that *z* is almost linear meaning thereby that *y* during this period is of the exponential type with

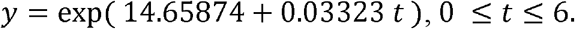

**Table-1:** First Order Differences of *z from* April 19 to April 25.

| Day | $Y$ | $Z$ | $\Delta z$ |
| --- | --- | --- | --- |
| April 25 | 2836588 | 14.85811 | 0.03250 |
| April 24 | 2745878 | 14.82561 | 0.03930 |
| April 23 | 2640059 | 14.78631 | 0.03355 |
| April 22 | 2552928 | 14.75275 | 0.03181 |
| April 21 | 2473002 | 14.72094 | 0.03090 |
| April 20 | 2397759 | 14.69004 | 0.03130 |
| April 19 | 2323875 | 14.65874 | 0.03326 |

Here 0.03323 is average of the values of Δ*z*.

Similarly, from Table-2, we can observe that *z* is almost linear and therefore *y* during this period is of the exponential type with

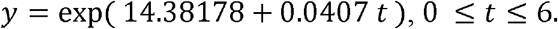

**Table-2:** First Order Differences of *z* from April 12 to April 18.

| Day | $Y$ | $Z$ | $\Delta z$ |
| --- | --- | --- | --- |
| April 18 | 2247861 | 14.62548 | 0.03701 |
| April 17 | 2166173 | 14.58847 | 0.04059 |
| April 16 | 2080001 | 14.54787 | 0.04434 |
| April 15 | 1989773 | 14.50353 | 0.04284 |
| April 14 | 1906336 | 14.46069 | 0.03932 |
| April 13 | 1832839 | 14.42137 | 0.03959 |
| April 12 | 1761700 | 14.38178 | 0.04179 |

We have mentioned earlier that the spread pattern is not a *monotonically* increasing function of time, and that trying to impose one monotone function for the entire available data is of no use. Indeed that is what has been reflected in Tables 1 and 2. It may be observed that the value of *b* in *y* = exp (*a* + *bt*) is in a decreasing trend with respect to time, which can be seen in the Tables 3, 4 and 5 also.

**Table-3:**
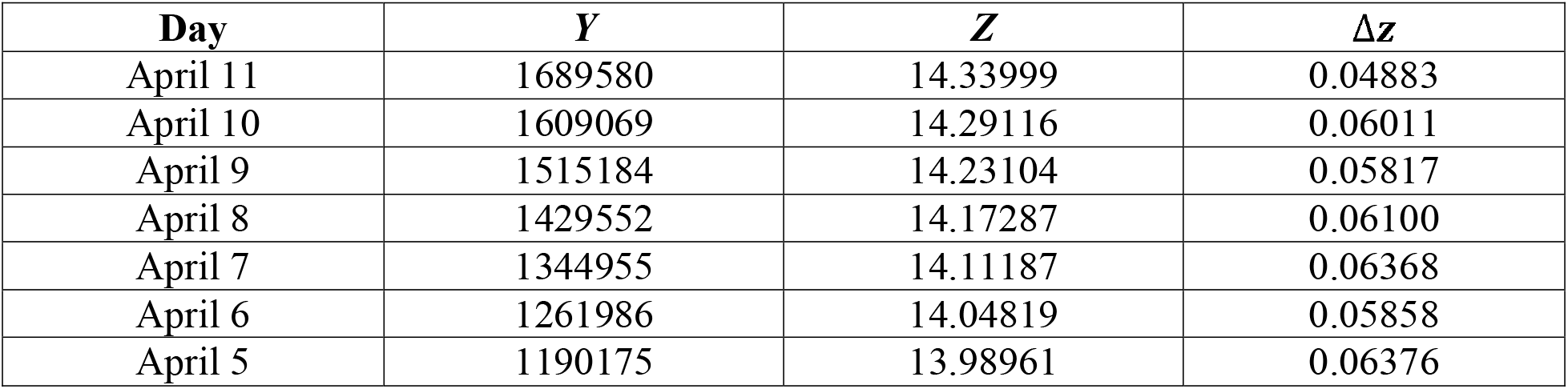
First Order Differences of *z* from April 5 to April 11.

**Table-4:**
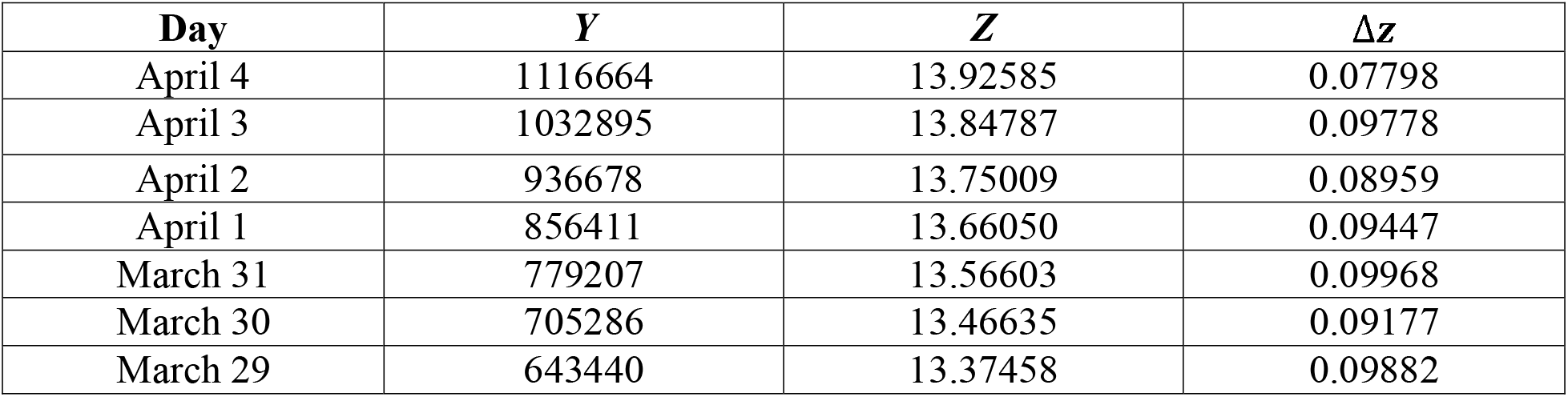
First Order Differences of *z* from March 29 to April 4.

**Table-5:** First Order Differences of *z* from March 21 to March 28.

| Day | $Y$ | $Z$ | $\Delta z$ |
| --- | --- | --- | --- |
| March 28 | 582898 | 13.27576 | 0.12178 |
| March 27 | 516063 | 13.15398 | 0.13372 |
| March 26 | 451469 | 13.02026 | 0.14522 |
| March 25 | 390444 | 12.87504 | 0.13278 |
| March 24 | 341898 | 12.74226 | 0.13719 |
| March 23 | 298067 | 12.60507 | 0.14975 |
| March 22 | 256614 | 12.45532 | 0.13525 |
| March 21 | 224151 | 12.32007 | 0.14073 |

From the tables, it may be observed that there are observable shifts in the values of Δ*z* the cut off dates being March 29, April 5, April 12 and April 19. From Table-3, we can observe that here too *z* is almost linear and therefore *y* during this period is nearly exponential with

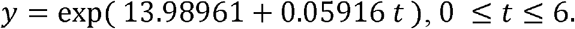

Table-4 also reflects a similar observation, and we get for that period of time

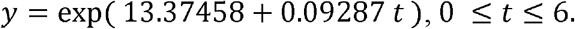

Finally, from Table-5 we get

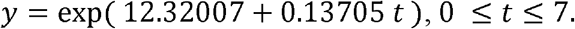

It may be seen that in Table-5 the values of Δ*z* are not quite following even approximate constancy. Therefore, we would not like to say that the data in Table-5 are of an exponential type, but it is true that though not exponential, the spread data in this period are very highly nonlinear. From the data, it becomes obvious that the data were nonlinear right from the start, i.e. from January 22. The nonlinearity increased from around March 7 onwards, and by March 29 (Table-4) it became nearly exponential.

## DISCUSSIONS

Researchers the world over have been working on mathematical modeling of COVID-19 data. It is but natural that different procedures of data analysis would be applied in the search for a proper mathematical model. Using classical mathematics easily understandable, we have tried to give a simple answer as to how the spread of the virus is expected to continue.

We have found that trying to fit a mathematical model defining the total number of COVID-19 cases as a monotonically increasing function of time for the entire data would not be a meaningful exercise because the dependent variable has not followed a monotone pattern. We have found that after around March 21, 2020, the spread pattern started to become nearly exponential. After March 28, it actually became of the exponential type but the pattern kept on changing after every seven days. So we cannot even say that even after March 28 one particular exponential curve would describe the dependent variable. We have seen that in

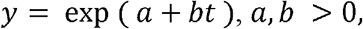

the coefficient *b* became equal to around

i. 0.13705 from March 21 to March 28,
ii. 0.09287 from March 29 to April 4,
iii. 0.05916 from April 5 to April 11,
iv. 0.04078 from April 12 to April 18, and
v. 0.03181 from April 19 to April 25.

In other words, the data are following approximately exponential patterns, and the patterns have been changing at almost regular intervals of seven days. Therefore trying to fit one single monotone function is of no use.

We would like to mention at this point that extrapolation using our simple method returns nearly accurate results. In Table-6, we have shown the expected values of the total cases outside China from April 26 to April 30. For the purpose of comparison we have shown the observed values also. The observed values during this period are from the Worldometer.info data published on May 1.

**Table-6:** Comparison of Forecasts and Observed Values.

| Day | $T$ | Observed $Y$ | Expected $Y$<br>(Forecasts) |
| --- | --- | --- | --- |
| April 26 | 1 | 2910465 | 2910455 |
| April 27 | 2 | 2977114 | 2986253 |
| April 28 | 3 | 3053669 | 3064025 |
| April 29 | 4 | 3135325 | 3143822 |
| April 30 | 5 | 3221358 | 3225698 |

As expected, on April 26, the value of Δ*z* became 0.02571 which is observably different from what it was in April 25. Now, using the value of Δ*z* = 0.02571, we have extrapolated the expected values from April 26 to April 30. From Table-6, it can be seen that our forecasts are quite near the observed values of the total number of cases.

We have observed that the values of Δ*z* are very slowly decreasing with time. Therefore for long term forecasting this method will give us a slightly overestimated value. For example in Table-6 above we have seen that the observed values and the forecasts are not too highly different. These forecasts were however for a short period of a maximum of five days. However, if we want to forecast what would be the approximate value of the total number of cases on May 23 for example, using Δ*z* = 0.02571, with April 25 as base, we would get a value that can be expected to be slightly more than the observed value on May 23.

Further, if the countries which have been contributing in observably large numbers towards the total number of cases outside China start to have diminishing trend in the total number of cases from some point of time, then also the forecast would be higher than the observed value. It is obvious that such a diminishing trend in any particular country is but expected. Therefore, if we use our model for long time forecasting, it would give a slightly overestimated value.

On an assumption that the total number of cases would keep on increasing at this rate, by May 23 the total number of COVID-19 cases outside China should be slightly less than 5.8 million. Indeed with April 25 as the base, for May 31 the value of *t* is 28. Using Δ*z* = 0.02571 which was used to calculate the expected value from April 26 to April 30, the expected value for May 23 is 5,826,866. It may be noted that for forecasting we have used a monotonically increasing function, although we have found that the pattern has been changing in an almost regular interval of 7 days.

It can be seen from the Worldometers.info data that the total number of cases outside China on May 23 is 5,314,979, as per their publication dated May 24. Our forecast with base as April 25 is 5,826,866. As expected, it is an overestimation. However if we would have considered recent data, of the second week of May for example, our forecast would have been much nearer the actual value. Hence although not quite fit for long term forecasts, our method can be used for short term forecasting.

One important point that we would like to point out is that the coefficient *b* is getting smaller and smaller with reference to time. It can be observed that while *b* has been decreasing, the parameter *a* has been consistently increasing. Therefore, even though *b* is getting smaller and smaller in time, the spread pattern is continuing to remain nearly exponential. The total number of cases on any given date may actually be more than what has been announced officially. Therefore an exact estimate of the pattern is not possible anyway.

The Geographical area, outside China, is heterogeneous. Therefore the spread pattern has to be different from region to region. In the totality, regional heterogeneity therefore is inherent. In India for example, as per the Worldometers.info data, COVID-19 cases were first detected on February 15, and this was the same day on which cases were first detected in the United States also. But as on April 25, the spread patterns in these two countries were very different. In our analysis, the total number of cases includes data from these two countries also which obviously are in two different stages of spread. Further, newer entries country-wise started to enter into the list at different times. So the cumulative total includes data from countries in different stages of spread. Still, we have found that there indeed is a nearly exponential type of spread pattern and that it is changing in about seven days or so. The way the spread is continuing the world over, it is clear the virus will continue to take its toll until medical science finally takes over.

## CONCLUSIONS

The spread curve of COVID-19 outside China was nonlinear right from the start. Since March 21, it is currently following nearly exponential patterns of growth with dependence on time changing at almost regular intervals of seven days. Unless the pandemic comes to a halt naturally or otherwise, the spread would grow enormously. Our simple method works quite well for short term forecasting.

## Data Availability

The data have been taken from Worldometers.info.

https://www.worldometers.info/coronavirus/coronavirus-cases/

## Notes

### Competing Interest Statement

The authors have declared no competing interest.

### Funding Statement

This work is not supported by any funding agency.

### Author Declarations

This is a mathematical study. This does not need any certification from any organization.

## REFERENCES

1. Worldometers.info. Total Cases excluding mainland China, Publishing Date: 30 April, 2020. Place of Publication: Dover, Delaware, U. S. A.

2. Ciufolini I, Paolozzi A. Mathematical prediction of the time evolution of the COVID-19 pandemic in Italy by Gauss error function and Monte Carlo Simulations. Eur. Phys. J. Plus. 2020;135:355. https://doi.org/10.1140/epjp/s1330-020-00383-y.

3. Kucharski AJ, Russel TW, Diamond C, et. al. Early dynamics of transmission and control of COVOD-19: a mathematical modeling study. The Lancet Infect. Dis. 2020;20(5):553–8. https://doi.org/10.1016/S1473-3099(20)30144-4.

4. Gondauri D, Mikautadze E, Batiashvilli M. Research on COVID-19 virus spreading statistics based on the examples of the cases from different countries. Elect. J. of Gen. Med. 2020;17(4). em209. https://doi.org/10.29333/ejgm/7869.

5. Wu JT, Leung K, Leung GM. Nowcasting and forecasting the potential domestic and international spread of the 2019 nCoV outbreak originating in Wuhan, China: A modeling study. The Lancet. 2020;395(10225):689–97. https://doi.org/10.1016/50140-6736(20)30260-9.

6. Baud D, Qi X, Nielson-Saines K, et. al. Real estimates of mortality following COVID-19 infection. The Lancet Infect. Dis. 2020. https://doi.org/10.1016/S1473-3099(20)30195-X.

7. Wang L, Li J, Guo S, et. al. Real-time estimation and prediction of mortality caused by COVID-19 with patient information based algorithm. Sc. of the Total Env. 2020;27. https://doi.org/10.1016/j.scitotenv.2020.138394.

